# The Mediating Role of Patient Satisfaction in the Relationship Between Healthcare Quality and Patient Loyalty in Health Insurance Hospitals in Alexandria, Egypt

**DOI:** 10.64898/2026.01.05.26343432

**Authors:** Doaa S. Galal, Magda R. Ahmed, Ekram W. Abd El-Wahab, Basem F. Abdel-Aziz

**Author notes:** To whom correspondence should be addressed: Ekram W. Abd El-Wahab, Department of Tropical Health, High Institute of Public Health, Alexandria University 165 El Horreya Road, 21561 Alexandria, Egypt.

## Abstract

**Background:** Healthcare quality is a key determinant of patient experience, satisfaction, and loyalty, yet the mechanisms linking quality to loyalty remain underexplored in Egypt’s Health Insurance Organization (HIO). This study examines the mediating role of patient satisfaction in the relationship between healthcare quality and patient loyalty among HIO beneficiaries.

**Methods:** A cross-sectional study was conducted with 983 beneficiaries from four HIO hospitals in Alexandria. Data was collected using a validated questionnaire covering eight healthcare-quality domains, patient satisfaction, and loyalty. Confirmatory factor analysis (CFA) assessed construct validity, and structural equation modeling (SEM) tested direct and mediated pathways. Bootstrapping (5,000 samples) evaluated mediation significance.

**Results:** Healthcare quality significantly predicted patient satisfaction (β = 0.607, p < 0.001), which in turn strongly influenced patient loyalty (β = 0.545, p < 0.001). Direct effects of healthcare quality on loyalty were reduced and non-significant when satisfaction was included. The indirect effect via patient satisfaction was significant (β = 0.331, 95% CI: 0.262–0.404), confirming partial mediation. Transition of Care, Empathy, and Responsiveness were the most influential quality domains. Model evaluation demonstrated a strong fit across all indices (CFI = 0.967, TLI = 0.960, RMSEA = 0.043).

**Conclusion:** Patient satisfaction is a key link between healthcare quality and loyalty. Enhancing communication, competence, responsiveness, and transitional care as satisfaction-driven domains can strengthen trust, continuity, and loyalty, supporting patient-centered reforms and Egypt’s Universal Health Insurance implementation.

**What is already known:**

- Healthcare quality strongly influences patient satisfaction and loyalty.
- Patient loyalty is critical for continuity of care, trust, and system sustainability.
- Few Egyptian studies have applied structural equation modeling to explore mediation pathways between quality, satisfaction, and loyalty.

**What this study adds:**

- Validation of a mediation model examining healthcare quality, patient satisfaction, and loyalty among HIO beneficiaries in Alexandria.
- Patient satisfaction partially mediates the relationship between healthcare quality and loyalty.
- Identifies key quality domains that drive satisfaction-related loyalty, providing actionable targets for improvement.

**How this study might affect research, practice, or policy:**

- Supports integrating patient satisfaction as a core indicator in HIO performance monitoring.
- Informs targeted quality-improvement interventions focusing on satisfaction-sensitive domains.
- Provides a robust mediation framework to guide evidence-based UHI policy and implementation in Egypt.

## INTRODUCTION

Patient loyalty, defined as a patient’s intention to revisit a healthcare provider and their willingness to recommend its services, is a critical indicator of health system performance and sustainability (1, 2). In publicly funded health systems, such as Egypt’s Health Insurance Organization (HIO), fostering loyalty is particularly important to minimize patient leakage to private providers, strengthen system utilization, and enhance continuity of care (3, 4). High patient loyalty contributes not only to consistent service utilization but also to overall public trust in healthcare reforms, which is vital for the successful implementation of Universal Health Insurance (UHI) in Egypt (5).

Healthcare quality is widely recognized as a primary determinant of patient loyalty. Quality encompasses both clinical effectiveness and the broader patient experience, including staff competence, responsiveness, empathy, communication, facility tangibles, and organizational processes (6–8). Theoretical frameworks in service quality and patient experience suggest that patient satisfaction functions as a mediating mechanism, translating perceptions of healthcare quality into loyalty-related behavioral intentions (9, 10). In other words, while high-quality services may directly influence patient loyalty, much of this effect is filtered through the patient’s subjective evaluation of their satisfaction with care. Empirical studies across different settings consistently demonstrate that patient satisfaction significantly mediates the relationship between perceived quality and loyalty, highlighting satisfaction as a critical lever for healthcare managers aiming to enhance engagement and retention (2, 11, 12).

Despite the growing international evidence, research on the mediating role of patient satisfaction in Egypt remains limited. Previous studies in Egyptian healthcare contexts have primarily focused on direct relationships between quality and satisfaction or loyalty, without exploring the indirect effects through advanced analytical approaches such as structural equation modeling (13, 14). This gap is particularly relevant for HIO hospitals, where understanding the pathways through which quality improvements translate into patient loyalty is essential for designing effective, evidence-based interventions aligned with national UHI reform goals.

Addressing this gap, the present study aims to examine the mediating effect of patient satisfaction on the relationship between healthcare quality and patient loyalty among HIO beneficiaries in Alexandria, Egypt. Specifically, it seeks to validate the measurement models for healthcare quality, patient satisfaction, and loyalty, and to assess the direct and indirect relationships between these constructs using SEM. Additionally, the study explores whether healthcare quality may mediate the relationship between patient satisfaction and loyalty, providing a comprehensive understanding of the complex interplay between these factors.

By identifying satisfaction as a key mediating mechanism, this research contributes to both theory and practice. It offers empirical evidence to support patient-centered interventions, such as enhancing communication, improving staff competence, optimizing care coordination, and strengthening facility processes, which can foster loyalty, improve trust in public healthcare, and ultimately support the objectives of Egypt’s UHI system (5, 15–17).

## METHODS

The detailed methodology underpinning the first part of this study will be published elsewhere and is therefore summarized here for completeness. In that earlier work, an analytical cross-sectional design was implemented across four HIO hospitals in Alexandria, Egypt, using a stratified random sampling approach to recruit beneficiaries attending inpatient, outpatient, emergency, and support service units. The sampling yielded 983 participants, exceeding the minimum SEM requirement and ensured stable parameter estimation. Data was collected through structured face-to-face interviews using a comprehensive questionnaire that included sociodemographic variables and the validated Arabic version of the National Patient Satisfaction Survey (NPSS). The NPSS captured key constructs including healthcare quality domains (tangibles, empathy, responsiveness, competence, information, availability, transition of care, organizational processes), patient satisfaction, and patient loyalty, across 87 items with established reliability. Confirmatory factor analysis (CFA) and SEM were applied to validate measurement constructs and explore the relationships among quality, satisfaction, and loyalty. Building on these previously established methods, the present analysis focuses specifically on the mediation and one-path structural models.

### Mediation Effect of Patient Satisfaction Between Healthcare Quality and Patient Loyalty

The mediation model (Figure S1), developed from the theoretical framework in Figure S2, assessed whether patient satisfaction mediates the relationship between healthcare quality and patient loyalty. The hypotheses tested were: i) H5(0): Patient satisfaction does not mediate the relationship between healthcare quality and patient loyalty, ii) H5(A): Patient satisfaction mediates the relationship between healthcare quality and patient loyalty.

Mediation was evaluated following the four recognized categories: full (indirect-only) mediation, partial mediation, complementary mediation, and competitive mediation, depending on the significance and direction of direct and indirect effects (18).

### One-Path Relationships Among Healthcare Quality, Patient Satisfaction, and Patient Loyalty

The one-path model (Figure S3), grounded in the theoretical model in Figure S4, examined the sequential influence among the three constructs. The hypotheses were: i) H6(0): Higher healthcare quality does not lead to higher patient satisfaction, and higher patient satisfaction does not result in greater patient loyalty. ii) H6(A): Higher healthcare quality leads to higher patient satisfaction, which consecutively leads to greater patient loyalty.

This model tested the linear causal chain predicted in the theoretical framework.

### Mediation Effect of Healthcare Quality Between Patient Satisfaction and Patient Loyalty

A second mediation model (Figure S5), derived from the theoretical model in Figure S6, examined whether healthcare quality mediates the association between patient satisfaction and patient loyalty. The hypotheses were: i) H7(0): Healthcare quality does not mediate the relationship between patient satisfaction and patient loyalty, ii) H7(A): Healthcare quality mediates the relationship between patient satisfaction and patient loyalty.

Together, these models enabled a comprehensive assessment of direct, indirect, and sequential pathways linking healthcare quality, satisfaction, and loyalty.

### Statistical Analysis

Descriptive statistics, comprising frequencies, percentages, means, and standard deviations, were used to describe participant characteristics and to summarize the distributions of all measured scales. Internal consistency reliability was assessed using standard reliability testing procedures [Cronbach’s alpha and Composite Reliability (CR)]. Confirmatory factor analysis was conducted to evaluate the measurement properties of healthcare quality, patient satisfaction, and patient loyalty constructs. Model fit was assessed using multiple indices, including the chi-square to degrees of freedom ratio (χ²/df), Comparative Fit Index (CFI), Tucker–Lewis Index (TLI), Incremental Fit Index (IFI), Normed Fit Index (NFI), Goodness-of-Fit Index (GFI), Root Mean Square Error of Approximation (RMSEA), Standardized Root Mean Square Residual (SRMR), and Root Mean Square Residual (RMR). SEM was subsequently employed to examine the hypothesized direct and indirect relationships among the three constructs. Mediation effects were tested using the nonparametric bootstrapping procedure with 5,000 resamples to generate bias-corrected confidence intervals for all indirect effects.

## RESULTS

### Characteristics of the study population

The detailed characteristics of the study population will be published elsewhere. Briefly, the sample was predominantly female (57.7%), largely urban, with a mean age of 45.1 ± 11.05 years. About half of the respondents were employed (52.4%) and had secondary or intermediate education (36.4%). A notable proportion reported inadequate income (42.8%). The majority were married (67.2%) with moderate family size of three to four members (50.3%). Over half of the participants reported chronic health conditions, and nearly half reported long-term utilization of health-insurance services for more than six years. most participants reporting moderate family size, secondary-level education, and long-term utilization of health-insurance services.

### Preliminary Observations

Although overall healthcare quality was mostly rated as moderate, satisfaction and loyalty levels were comparatively higher, with nearly half expressing good or very good satisfaction and about 40% reporting strong loyalty. All measurement scales demonstrated high internal consistency, and the healthcare quality domains showed strong factor loadings and good model fit. After removing low-loading items and refining the measurement models, all factor loadings exceeded 0.7, and reliability, convergent validity, and discriminant validity were confirmed across constructs. Structural equation modeling indicated that healthcare quality significantly influenced its eight component dimensions, with the strongest effects observed for Transition of Care and Empathy. Healthcare quality also strongly predicted patient satisfaction and moderately predicted loyalty, while satisfaction substantially predicted loyalty. Total effects showed that improvements in healthcare quality produced significant increases in both satisfaction and loyalty. However, when tested as a mediation pathway, healthcare quality no longer had a significant direct effect on loyalty but retained a significant indirect effect through satisfaction, confirming satisfaction as a key mediator in the relationship between healthcare quality and patient loyalty.

### Mediation of Patient Satisfaction Between Healthcare Quality and Patient Loyalty

Table 1 presents the assessment of the mediating effect of patient satisfaction on the relationship between HC quality and patient loyalty. The model demonstrated excellent fit, with χ² (CMIN) = 266.676, df = 101, p < 0.001, and a χ²/df ratio of 2.64. Additional fit indices were within acceptable to excellent ranges: Root Mean Square Residual (RMR) = 0.048, Goodness-of-Fit Index (GFI) = 0.964, Normed Fit Index (NFI) = 0.948, Incremental Fit Index (IFI) = 0.967, Comparative Fit Index (CFI) = 0.967, and Tucker–Lewis Index (TLI) = 0.960. The Root Mean Square Error of Approximation (RMSEA) was 0.043, with PCLOSE = 0.967, and the Akaike Information Criterion (AIC) = 336.676.

**Table 1:** Assessment of the mediating effect of patient satisfaction on the relationship between HC quality and patient loyalty.

| Path/Relationship | Standardized estimation | Direct effect | Indirect effect | Confidence interval |  | Significance | Significance Impact |
| --- | --- | --- | --- | --- | --- | --- | --- |
|  |  |  |  | Lower Bound | Upper Bound |  |  |
| Health Care quality → patient satisfaction → patient loyalty | 0.389 | 0.814 | 1.2362 | 0.609 | 2.341 | 0.004 | Full mediation |

The structural paths showed that HC quality did not significantly predict patient loyalty (B = 0.814, SE = 0.523, CR = 1.558, p = 0.119), leading to acceptance of the null hypothesis for this relationship. In contrast, HC quality significantly predicted patient satisfaction (B = 0.609, SE = 0.244, CR = 2.501, p = 0.012). Patient satisfaction also significantly predicted patient loyalty, supporting its role as an important pathway between HC quality and loyalty.

### Direct and Indirect Effects Between Healthcare Quality, Patient Satisfaction, and Patient Loyalty

The direct and indirect effects were examined through bootstrapping. The one-path model showed that HC quality exerted a strong direct effect on patient satisfaction, which in turn influenced loyalty. The standardized indirect effect (HC quality → patient satisfaction → patient loyalty) was 1.034. The direct effect was 2.464, and the indirect effect was 2.494. The bootstrapped confidence interval for the indirect effect ranged from 1.703 to 2.494, with a p-value of 0.009, indicating a highly significant mediating effect (p < 0.01). These results support the alternative hypothesis (H6), confirming that higher HC quality leads to increased patient satisfaction, which subsequently enhances patient loyalty (Table 2).

**Table 2:** Full mediation effect of Patient Satisfaction on the relationship between healthcare quality and Patient Loyalty.

| Path/Relationship | Standardized estimation | Direct effect | Indirect effect | Confidence interval |  | Significance | Significance Impact |
| --- | --- | --- | --- | --- | --- | --- | --- |
|  |  |  |  | Lower Bound | Upper Bound |  |  |
| Satisfaction →<br>Health Care<br>quality → Loyalty | 1.034 | 2.464 | 2.494 | 1.703 | 2.494 | 0.009 | highly significant |

### Mediating Effect of Healthcare Quality on the Relationship Between Patient Satisfaction and Patient Loyalty

Table 3 evaluates whether HC quality mediates the relationship between patient satisfaction and loyalty. Patient satisfaction had a strong direct effect on HC quality (B = 0.962, SE = 0.039, CR = 11.562, p < 0.001; CI: 0.378–0.533). However, HC quality did not significantly predict patient loyalty (B = 0.389, SE = 0.523, CR = 1.558, p = 0.119; CI: –0.211 to 1.839). In contrast, patient satisfaction significantly predicted loyalty (B = 0.613, SE = 0.244, CR = 2.501, p < 0.001; CI: 0.130–1.087).

**Table 3:** The mediating effect of healthcare quality on the relationship between patient satisfaction and loyalty.

3: The mediating effect of healthcare quality on the relationship between patient satisfaction and loyalty.
| Path/Relationship | Standardized estimation | Direct effect | Indirect effect | Confidence interval |  | Significance | Significance Impact |
| --- | --- | --- | --- | --- | --- | --- | --- |
|  |  |  |  | Lower Bound | Upper Bound |  |  |
| Patient satisfaction →<br>Health Care quality →<br>patient loyalty | 0.613 | 0.609 | 0.371 | 0.008 | 1.709 | 0.087 | No Mediation |

Total effects showed that patient satisfaction influenced HC quality (0.456) and loyalty (0.981), and that HC quality had a total effect of 0.814 on loyalty. The indirect effect of patient satisfaction on loyalty through HC quality was 0.371. Bootstrapping yielded p = 0.087 and a confidence interval of 0.008–1.709. Because the indirect effect was not statistically significant at p < 0.05, the null hypothesis was accepted, indicating that HC quality does not significantly mediate the relationship between patient satisfaction and patient loyalty.

#### Measurement Model (CFA)

The results of CFA demonstrated strong construct validity. All factor loadings exceeded 0.70. Reliability indicators as included by Cronbach’s alpha and Composite Reliability (CR), were above 0.80. Average Variance Extracted (AVE) values exceeded the recommended threshold of 0.50. Overall model fit was strong (CFI = 0.943, TLI = 0.940, RMSEA = 0.031).

#### Structural Model (Direct Effects)

Healthcare quality had a strong positive effect on patient satisfaction (β = 0.607, p < 0.001) and a moderate but significant effect on patient loyalty (β = 0.263, p < 0.001). Patient satisfaction exhibited a strong and significant effect on loyalty (β = 0.545, p < 0.001).

#### Mediation Analysis

Bootstrapping confirmed that patient satisfaction partially mediated the relationship between healthcare quality and patient loyalty. The indirect effect was significant (β = 0.331, 95% CI: 0.262–0.404). The direct effect remained significant but reduced in magnitude, indicating partial-not full-mediation.

#### Model Fit Indices

Figure 1 presents the theoretical model assessing the direct relationships among healthcare quality, patient satisfaction, and patient loyalty.

**Figure 1:**
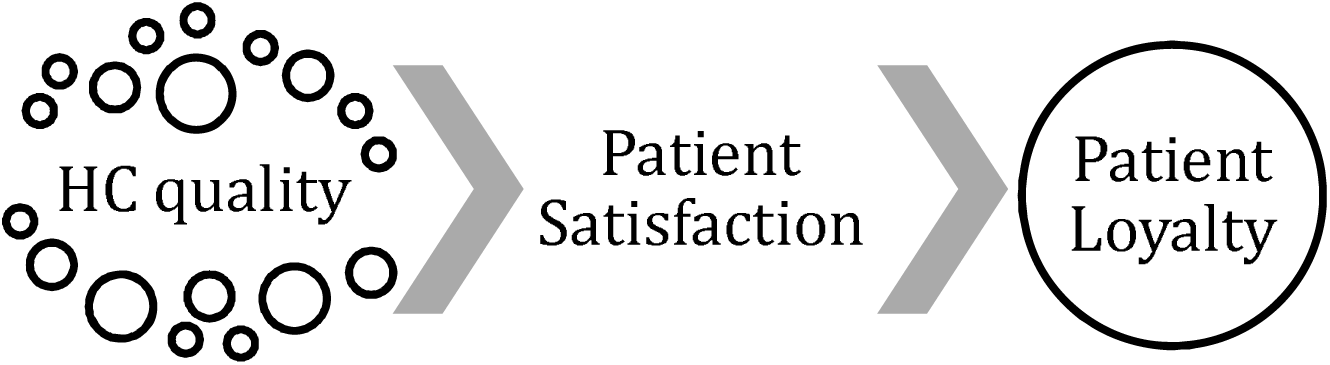
Theoretical model for a direct relationship between the Quality of Health Care, Patient Satisfaction, and Loyalty.

#### SEM Analysis for Mediation Hypothesis Testing

- Patient Satisfaction as a Mediator Between Healthcare Quality and Patient Loyalty The model demonstrated excellent fit: χ² = 266.676, df = 101, p < 0.001; χ²/df = 2.640; RMR = 0.048; GFI = 0.964; NFI = 0.948; IFI = 0.967; CFI = 0.966; TLI = 0.960; RMSEA = 0.043; PCLOSE = 0.967; AIC = 336.676 (Figure 2).
- One-Path Relationship Among Healthcare Quality, Patient Satisfaction, and Patient Loyalty Figure 3 demonstrates a well-fitting model (χ² = 268.7, df = 102, p < 0.001; χ²/df = 2.635; RMR = 0.049; GFI = 0.964; NFI = 0.947; IFI = 0.967; CFI = 0.966; TLI = 0.960; RMSEA = 0.043; PCLOSE = 0.969). In this model, HC quality significantly influenced patient loyalty (B = 0.972, SE = 0.180, CR = 11.350, p < 0.01; CI: 1.6932–2.3988) and patient satisfaction (B = 0.944, SE = 0.047, CR = 21.207, p < 0.01; CI: 0.897–1.081).
- Healthcare Quality as a Mediator Between Patient Satisfaction and Patient Loyalty Fit indices again indicated strong fit: χ² = 266.676, df = 101, p < 0.001; χ²/df = 2.640; RMR = 0.048; GFI = 0.964; NFI = 0.948; IFI = 0.967; CFI = 0.967; TLI = 0.960; RMSEA = 0.043; PCLOSE = 0.967; AIC = 336.676 (Figure 4).

## DISCUSSION

Healthcare quality significantly influences patient satisfaction and loyalty among public insurance beneficiaries. By integrating eight domains; tangibles, empathy, responsiveness, competence, information and communication, availability of services, organizational processes, and transitions of care, the validated model demonstrates that healthcare quality is multidimensional, encompassing both interpersonal interactions and system-level processes that ensure continuity, coordination, and accessibility. Structural analyses confirmed that improvements across these domains increase patient satisfaction, which in turn mediates the relationship between quality and loyalty. While quality alone does not directly predict loyalty, satisfaction emerges as the key determinant of patient commitment, echoing global findings across health systems.

**Figure 2:**
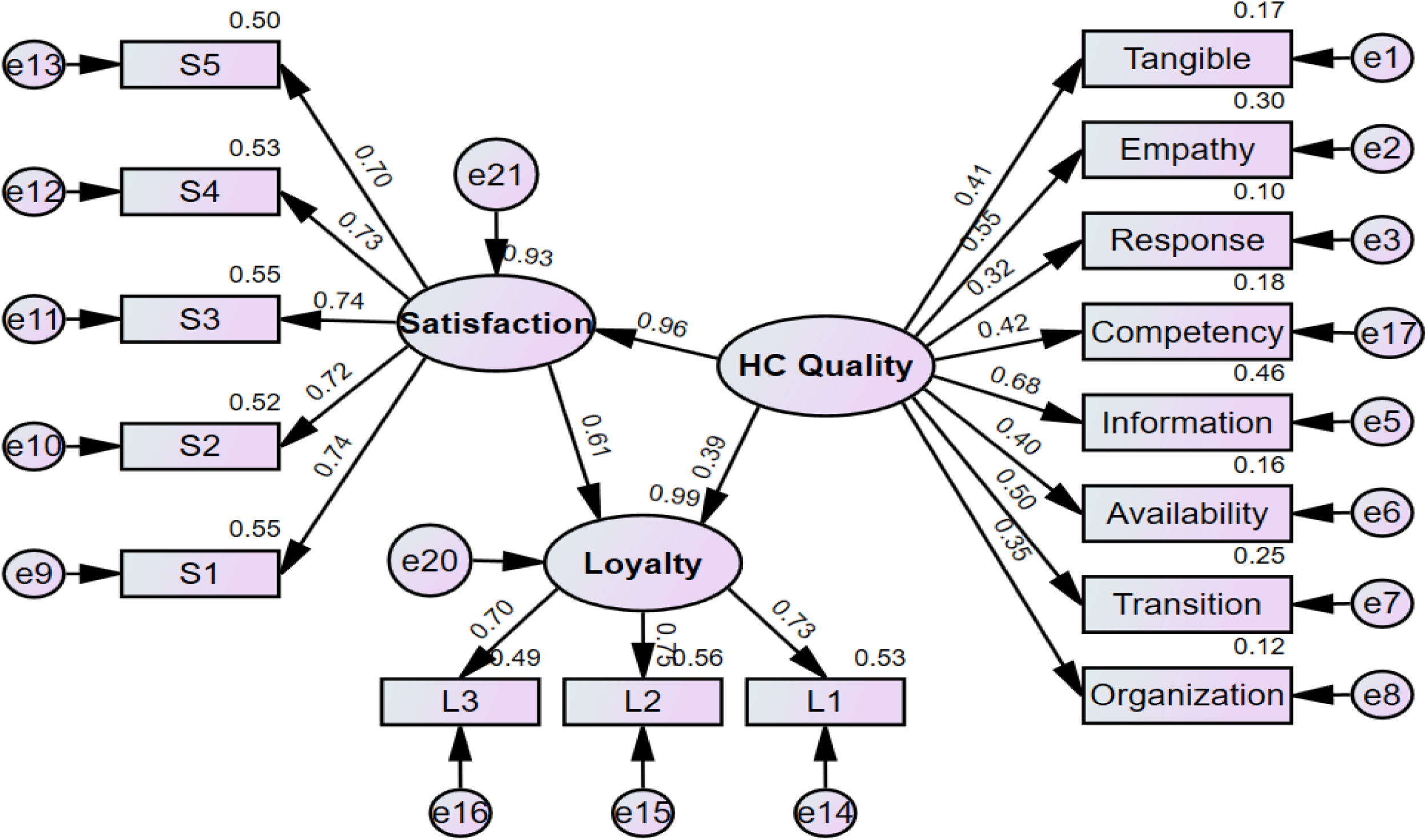
Patient satisfaction mediates the relationship between Health Care quality and patient loyalty.

**Figure 3:**
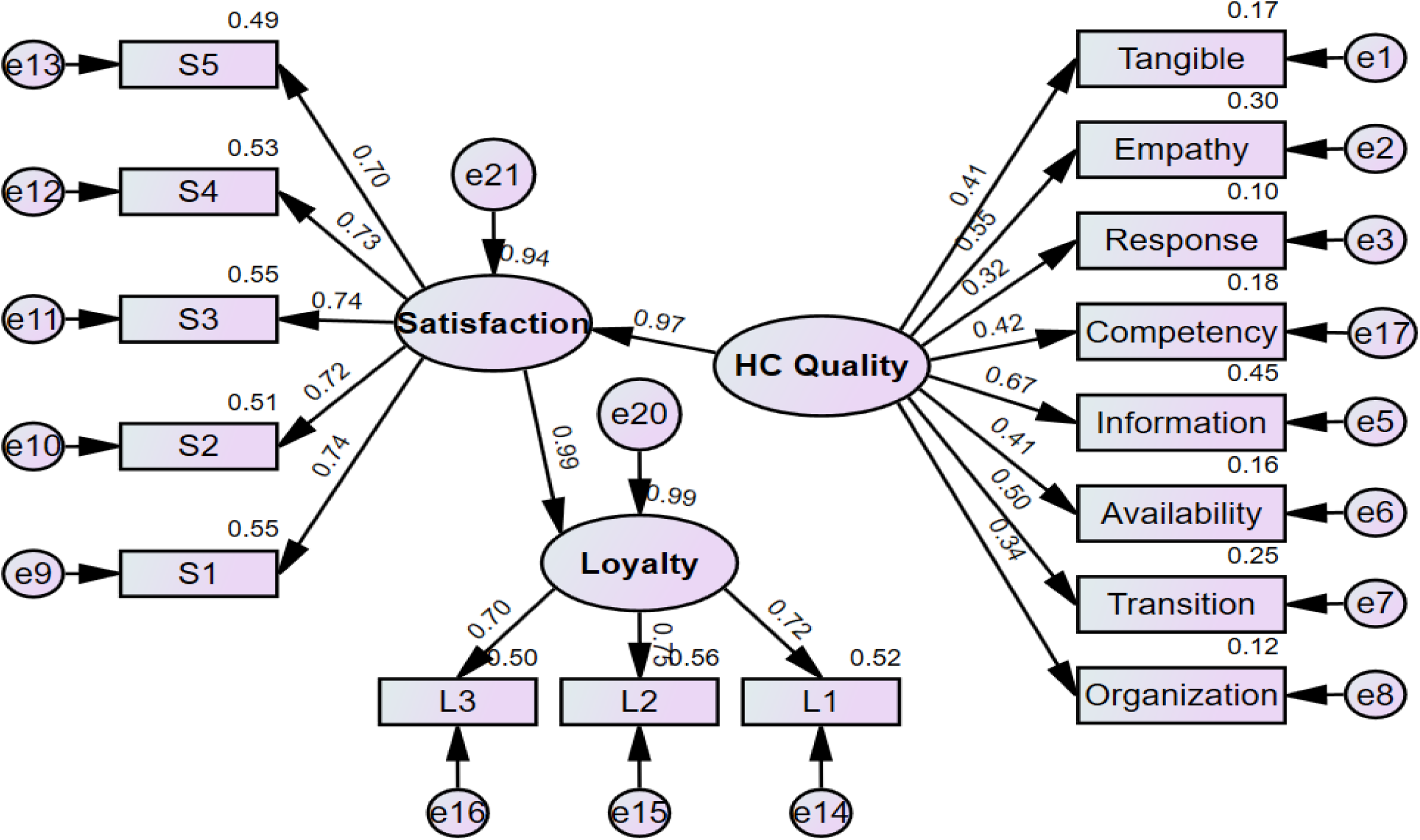
Theoretical model of the one-path relationships between Health Care quality, patient satisfaction, and patient loyalty.

**Figure 4:**
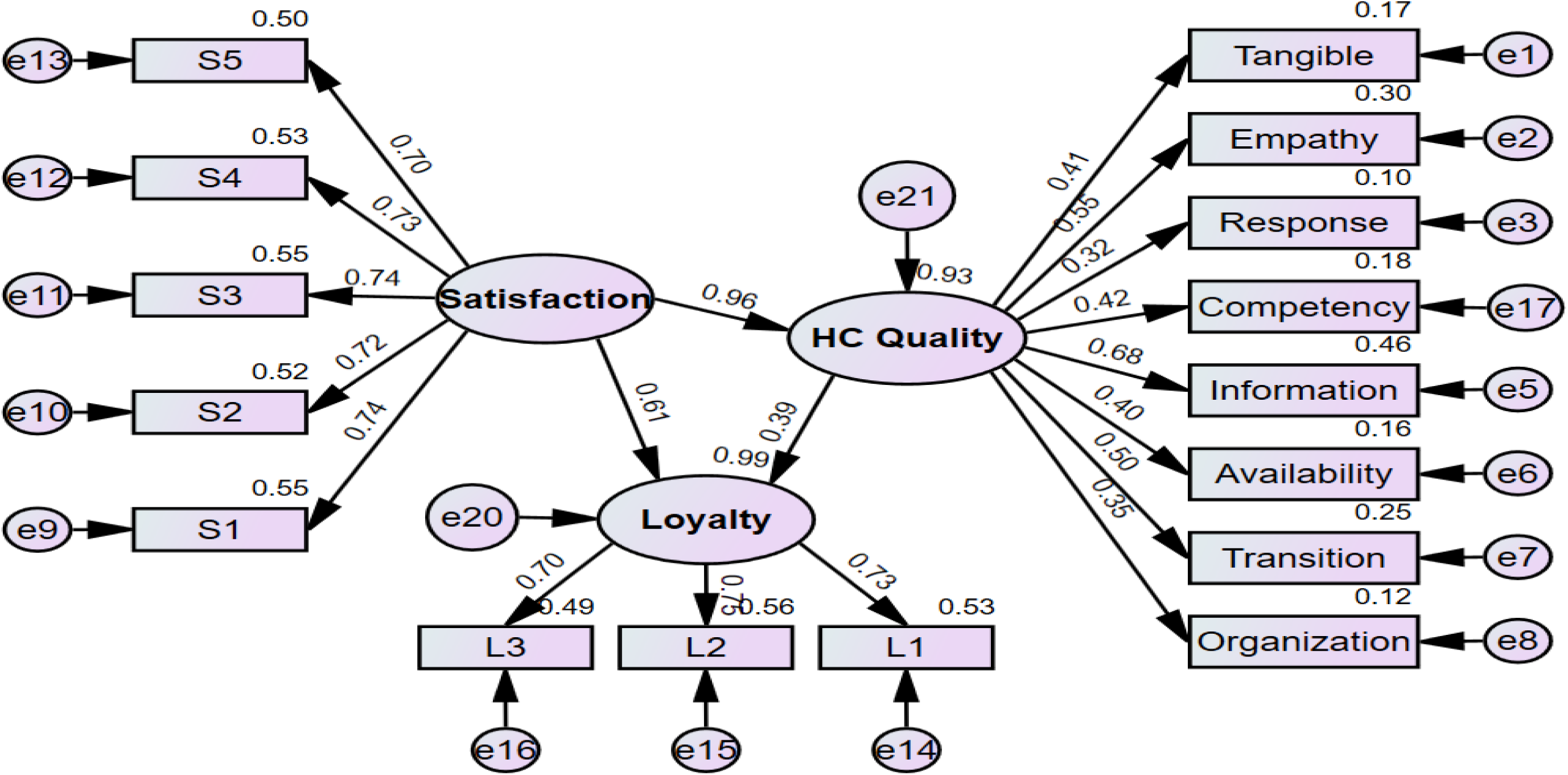
Health Care quality mediated the relationship between patient satisfaction and patient loyalty.

The results have critical implications for public insurance contexts, where loyalty supports continuity of care, efficient resource utilization, and system sustainability. Enhancing communication, care coordination, staff competence, and facility organization is essential for fostering trust, adherence, and repeated service use. By adopting this holistic, eight-domain quality framework, policymakers and healthcare leaders can guide patient-centered reforms, strengthen satisfaction and loyalty, and promote equitable, sustainable engagement with public healthcare services, supporting broader universal health insurance objectives.

While healthcare quality alone does not directly predict loyalty, satisfaction emerges as the central determinant of patient commitment, highlighting the importance of patient-centered interventions that foster trust, continuity of care, and repeated engagement with public healthcare services (6–8). These results highlight that patients evaluate healthcare holistically, considering not only the technical competence of providers but also interpersonal interactions, organizational efficiency, and system-level processes that support accessibility and continuity.

Many previous studies align with these findings, that patient satisfaction significantly mediates the relationship between healthcare quality and patient loyalty. Consistently, satisfaction strongly affects both perceived quality and loyalty, while the direct effect of quality on loyalty is minimal or non-significant. Research in outpatient and hospital contexts confirms that satisfaction fully or partially mediates the link between service quality and loyalty, emphasizing that patients commit to providers primarily when their expectations and experiences are met (3, 4, 9, 11). Similarly, studies in Jordan and other health systems show that patient satisfaction serves as a significant mediator in the relationship between service quality dimensions and loyalty, reinforcing the universal relevance of patient-centered care (14).

However, some research presents contrasting evidence. Liu and colleagues found that patient satisfaction did not mediate the relationship between healthcare quality dimensions and loyalty, suggesting that contextual factors such as patient expectations, cultural norms, and the structure of healthcare systems can influence the strength of these relationships (13, 19–21). These discrepancies highlight the need to consider both organizational and socio-cultural contexts when designing quality improvement strategies, as the same interventions may produce varying outcomes across different healthcare environments.

The present study emphasizes that healthcare quality is not only a driver of satisfaction but must be understood as a comprehensive construct encompassing clinical competence, organizational efficiency, communication, and patient experience. Improvements in these domains consistently lead to higher satisfaction, which then translates into greater loyalty. This has significant implications for public health insurance systems, where patient loyalty contributes to continuity of care, system utilization, and the sustainability of reforms. Investments in staff training, standardized communication protocols, workflow optimization, and structured transitional care can therefore enhance satisfaction and promote repeated engagement with public healthcare facilities.

### Policy and practice implications

The findings of the present work highlight the necessity for public healthcare organizations to adopt holistic, patient-centered strategies targeting both clinical and non-clinical aspects of care.

Enhancing staff competence, communication, responsiveness, facility organization, and transitions of care can strengthen patient satisfaction and loyalty. Regular monitoring of patient experiences through surveys, feedback mechanisms, and benchmarking against best practices can guide continuous improvement. These measures not only improve individual patient outcomes but also support broader health system goals, including equitable access, continuity of care, trust in public healthcare services, and long-term sustainability of public insurance reforms (6–8).

In conclusion, patient satisfaction is a significant mediator between healthcare quality and loyalty. Targeted improvements in quality domains that influence satisfaction, such as staff competence, communication, responsiveness, and care transitions can meaningfully enhance patient loyalty and support the successful implementation of Egypt’s Universal Health Insurance reforms. These findings emphasize the importance of sustained investment in patient-centered quality improvement as a cornerstone of health system strengthening and long-term sustainability.

## Ethical consideration

### Ethical approval and consent to participate

The study was approved by the institutional review board and the ethics committee of the High Institute of Public Health affiliated with Alexandria University, Egypt. We sought the permission and support of the local health authorities to conduct the study in the selected districts in Alexandria. The study was conducted in accordance with the international ethical guidelines and of the Declaration of Helsinki. Informed written consent was obtained from each participant after explaining the aim and concerns of the study. Data sheets were coded by number to ensure anonymity and confidentiality of the participants’ data.

This article does not contain any studies with animals performed by any of the authors.

- **Informed consent:** Declared
- **Funding:** None
- **Conflicts of interest:** None to declare.

### Consent for publication

All authors approved the manuscript for publication

### Availability of supporting data

All data are fully available without restriction by the corresponding author at

## Data Availability

All data produced in the present study are available upon reasonable request to the authors

## Acknowledgements

We would like to acknowledge the study participants for accepting to participate in the study.

