## Supplementary material for "The Mediating Role of Patient Satisfaction in the Relationship Between Healthcare Quality and Patient Loyalty in Health Insurance Hospitals in Alexandria, Egypt": Suppl Figures

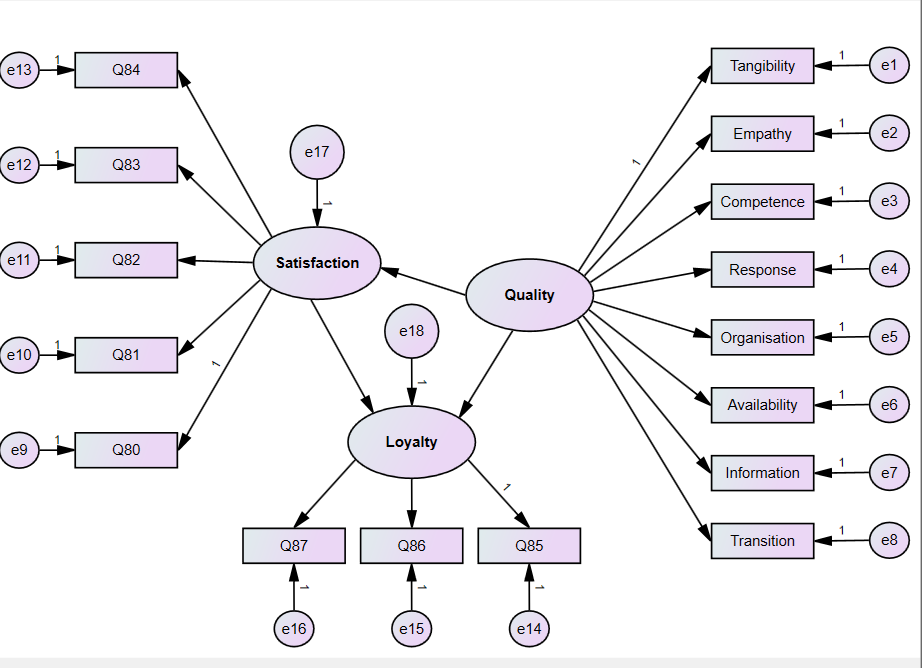

Figure S1: Study model of the mediation effect of patient satisfaction on the relationships between the Quality of Health Care and Patient Loyalty

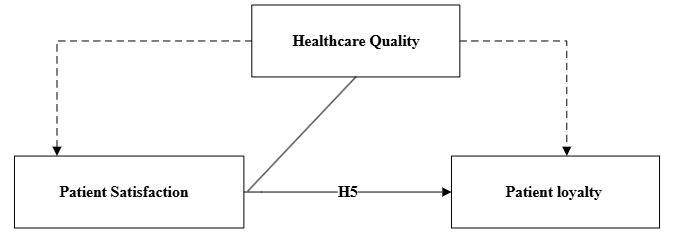

Figure S2: Theoretical model of the mediation effect of patient satisfaction on the relationships between Health Care Quality and Patient Loyalty

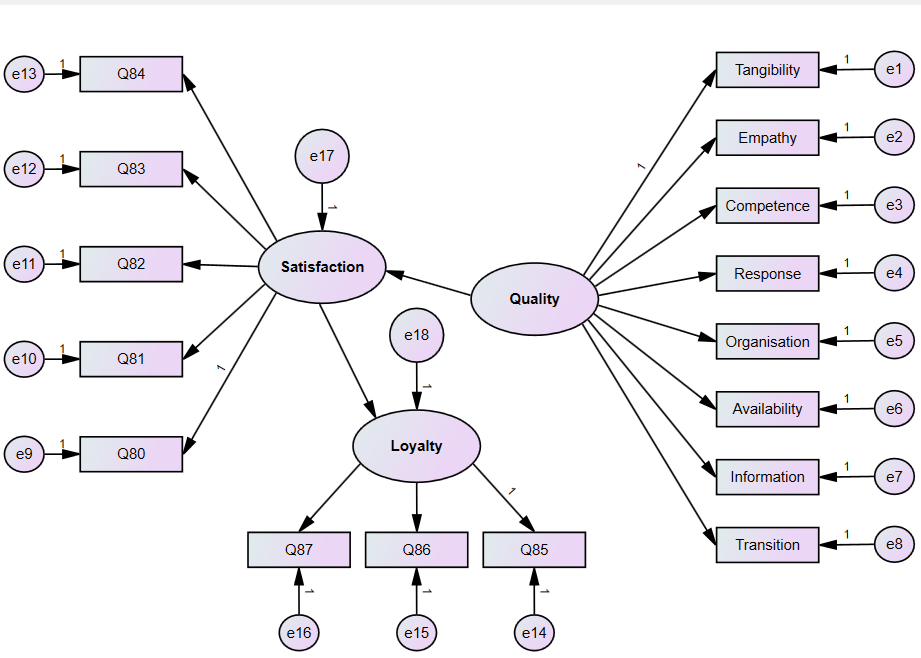

Figure S3: Study Model of the one-path relationships between Health Care quality, patient satisfaction, and patient loyalty

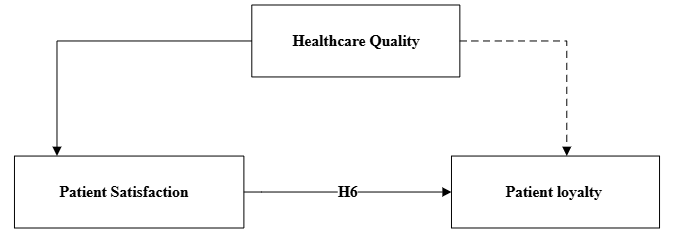

Figure S4: Theoretical model of the one-path relationships between Health Care quality, patient satisfaction, and patient loyalty

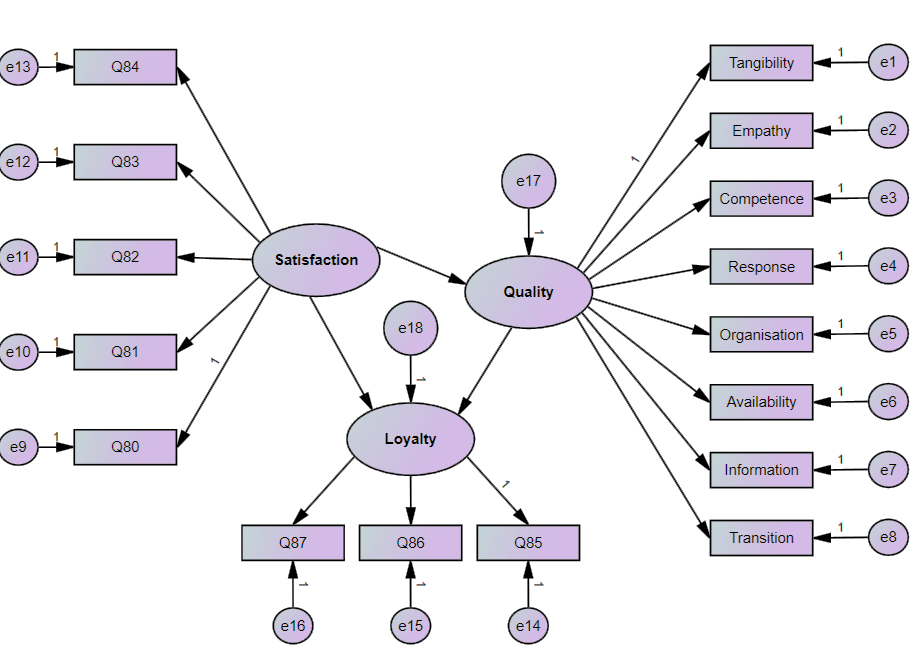

Figure S5: Study model of the mediation effect of Health Care Quality on the relationship between patient satisfaction and Patient Loyalty

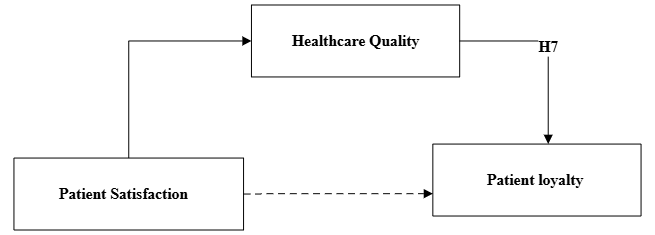

Figure S6: Theoretical model of the mediation effect of Health Care Quality on the relationships between patient satisfaction and Patient Loyalty
